# The effects of different course assessment methods on college students’ tennis performance and basic psychological needs: A cluster randomized controlled trial

**DOI:** 10.64898/2026.04.16.26351011

**Authors:** Yazhuo Wang, Yan Luo

**Author notes:** Corresponding author: Yazhuo Wang.

## Abstract

**Purpose:** This study aimed to examine the effects of formative and summative assessments on college students’ tennis performance and basic psychological needs.

**Methods:** A total of 128 undergraduate students (64 males, 64 females; *M*_age_ = 19.22, *SD* = 0.91) participated in this study. Participants were cluster-randomized to either a formative assessment group (*n* = 64) or a summative assessment group (*n* = 64). The formative assessment intervention involved setting personalized learning goals and success criteria, administering periodic tests, and providing process-oriented and individualized feedback. The summative assessment intervention involved setting uniform goals for all students, offering instructor feedback only on common problems, and requiring students to practice independently after class without personalized guidance. Both interventions were implemented over 10 weeks, with one 90-minute session each week. Tennis skills and basic psychological needs (i.e., autonomy, competence, and relatedness) were assessed before and after the intervention. Tennis skills were reassessed 1 week after the intervention. Two-way mixed-effects analysis of variance (ANOVA) was used to examine the impact of group, time, and their interaction on tennis skills and basic psychological needs.

**Results:** The results showed that the interaction between group and time was significant for all of the outcome variables. Simple effects analyses indicated that, at pre-test, the two groups did not differ significantly in tennis performance or in satisfaction of autonomy, competence, and relatedness (*p* > 0.05). At post-intervention, the formative assessment group demonstrated significantly better performance than the summative assessment group in tennis skills (*MD* = 3.50, 95% *CI* = [1.303, 5.697], *p* = 0.002), autonomy (*MD* = 2.44, 95% *CI* = [1.816, 3.059], *p* < 0.001), relatedness (*MD* = 1.33, 95% *CI* = [0.679, 1.977], *p* < 0.001), and competence (*MD* = 1.75, 95% *CI* = [1.046, 2.454], *p* < 0.001). At the 1-week follow-up session, the formative assessment group also showed significantly better tennis performance than the summative assessment group (*MD* = 6.81, 95% *CI* = [4.667, 8.958], *p* < 0.001).

**Conclusion:** Formative assessment was more effective than summative assessment in improving college students’ tennis performance and satisfying their basic psychological needs. These findings suggest that incorporating personalized goals, process-oriented evaluation, and individualized feedback into tennis instruction could promote both skill development and psychological outcomes in college physical education.

## Introduction

Empirical evidence revealed that sedentary behavior had become increasingly prevalent among college students. Data from self-reports found a daily sitting time of over eight hours, which was associated with significant detriment to both physical and mental health [1]. Tennis is effective in reducing sedentary behavior and improving physical and mental health. Research indicates that regular tennis activities can significantly reduce body fat percentage in adolescents and adults, lower the risk of heart disease, and enhance musculoskeletal (MSK) function and balance, thereby improving quality of life [2–5]. Furthermore, regular tennis participation helps alleviate symptoms of anxiety and depression, enhances cognitive flexibility, and promotes positive emotions [6, 7]. At the same time, this sport offers distinct advantages in fostering positive social interactions and strengthening social connections, and is therefore regarded as a preferred form of exercise that effectively supports health across the entire life span [8]. However, as its nature as an open-ended motor skill [9], the technical mastery of tennis relies heavily on continuous movement correction and immediate feedback during the learning process, such as coordinating force generation through the kinetic chain and judging the timing of shots [10]. Although regular tennis activities help improve cardiovascular health, promote musculoskeletal health, physical coordination, and neurocognitive function [3–5], traditional summative assessments of tennis no longer meet college students’ needs for autonomy, competence, and relatedness, thereby significantly diminishing their enthusiasm for participation and motivation to continue playing.

In recent years, the International Association for Higher Education in Physical Education [11] has recognized the importance of formative assessment in physical education and advocated for its implementation. Chinese physical education curriculum is also undergoing a paradigm shift from outcome-oriented “summative assessment” to process-oriented “formative assessment.” This transition is grounded in a “learner-centered” educational philosophy, which promotes students’ autonomous learning abilities and stimulates their intrinsic motivation and sustained participation in physical activity through strategies such as goal-setting and process-based evaluation [12–14]. Against this backdrop, tennis, a sport highly favored by college students, faces new opportunities and challenges in its teaching practices of Physical Education.

Physical education is widely recognized as the optimal setting for promoting the development of motor skills [2]. Common assessment strategies in physical education include formative and summative assessment. Summative assessment refers to a systematic evaluation conducted after the conclusion of a teaching phase to determine students’ learning outcomes or the extent to which predetermined objectives have been achieved [15]. Its primary purpose is to assign grades, certify competencies, or summarize learning outcomes, rather than to directly guide the immediate teaching process [16, 17]. While summative assessment can identify learning outcomes, it struggles to provide detailed feedback that promotes subsequent learning. Additionally, its “high-stakes” nature can easily cause students to feel significant pressure and anxiety [18–21]. In contrast, formative assessment focuses more on helping students understand their current level of learning and clarify expected learning goals [15]. It emphasizes that teachers evaluate students’ learning immediately, dynamically, and repeatedly during the teaching process, with a focus on timely feedback to reinforce and improve student learning. Common tools for formative assessment include self-assessment, peer assessment, learning journals, surveys, interviews, interim tests, and classroom observations [13]. The feedback generated by these assessment tools helps learners build confidence and motivation, enhance their reflective abilities, improve learning strategies, and better engage in monitoring their own learning processes [20, 22]. Research indicates that formative assessment effectively activates and supports students’ self-regulation abilities by continuously monitoring their learning progress and providing timely feedback [23, 24], making it a key strategy for promoting deep learning and self-regulation [25]. Compared to other educational interventions, formative assessment is more effective in enhancing teaching quality and promoting positive learning outcomes [13, 26]. In the field of physical education, formative assessment is also regarded as a teaching strategy with broad application prospects. Research indicates that integrating formative assessment into instruction can improve the effectiveness of elementary school physical education [17, 27]. Particularly in specific motor skill learning contexts, formative assessment practices, such as the use of digital video tools, reference to scoring rubrics, and guiding students in self-assessment, have been shown to significantly enhance students’ motivation and engagement, and to have a direct positive impact on their skill performance [28–30]. When formative assessment is integrated with specific sports education models, such as, teaching formats that incorporate real-game scenarios or tactical questioning, it can promote students’ motor skills and tactical decision-making abilities [31, 32].

Although physical education teachers place great importance on formative assessment [33], its implementation in teaching practice is fraught with difficulties [16, 26, 34, 35]. The challenges in implementing formative assessments can be broadly categorized into two types: individual factors and environmental factors. As decision-makers in teaching activities, teachers’ beliefs, knowledge base, and skill levels constitute the individual factors that determine the quality of formative assessment implementation [36, 37]. Secondly, teachers’ practical behaviors are heavily constrained by complex teaching contexts [38, 39]. For instance, the dynamic nature of classroom atmospheres and the unpredictability of student responses increase the difficulty of implementation [40]. Structural factors, such as internal school policies, particularly within an educational context dominated by summative assessment [41], may either grant teachers’ autonomy or restrict the flexibility of their assessment practices [42]. In short, there is little evidence that physical education teachers frequently use formative assessment strategies in their teaching [16]. Therefore, to better implement formative assessment, it is crucial to explore specific strategies for its design and implementation [17, 43].

However, there remains a significant gap in current empirical research on the practice of formative assessment in college tennis courses. Firstly, research have proved that formative assessment plays a positive role in general physical education [17, 27, 43], such as basketball, track and field, and volleyball [31, 32]. However, tennis is an open-ended motor skill involving complex movements and a high level of coordination, requiring continuous, diverse, and supportive feedback loops to help learners maintain focus and enhance their cognitive and behavioral performance in motor skills. Yet, applied research in this area remains extremely scarce. Secondly, existing evaluative studies have primarily focused on children or adolescents [17, 27, 29, 44, 45], while research targeting colleges students remains relatively scarce. Given the established risks of pervasive sedentary behavior and the promise of tennis as an intervention, a key unanswered question is the effect of formative assessment on beginners’ tennis performance. Investigating this is imperative to develop targeted, evidence-based strategies that mitigate health risks and foster student development. Consequently, the specific impact of formative assessment on the tennis performance of beginner college students has not yet been fully explored. Finally, research generally indicates that physical education teachers face numerous challenges in implementing formative assessments in practice, such as insufficient planning time, a lack of examples, and a culture dominated by traditional summative assessment [16, 34, 39, 43]. It remains unclear how these challenges will manifest and be addressed specifically within the context of teaching tennis to college students. Therefore, this study aims to explore the impact of different assessment methods on the tennis skills of college student beginners by designing and implementing formative and summative assessments in a real-world teaching context.

Basic Psychological Needs Theory (BPNT) provides an important theoretical framework for identifying environmental characteristics that promote individual behavioral motivation in physical education [44, 45]. BPNT is a core component of Self-Determination Theory (SDT) [46]and emphasizes that humans possess three basic psychological needs: autonomy, competence, and relatedness. These needs are innate to humans, universally present across cultures, and play a crucial role in individual growth, development, and health [47]. When the physical education environment satisfies these three needs, it promotes the internalization of extrinsic motivation, facilitates the formation of intrinsic goal orientation [48], and thereby enhances athletic performance [49, 50] and behavioral persistence [51]. The practical characteristics of formative assessment, such as supporting students’ autonomy, relatedness, and competence by providing choices, process feedback, and opportunities for collaboration, align closely with the core tenets of the BPNT. Although Herrero-González et al. [33] and Leenknecht et al. [14] suggest that formative assessment may enhance intrinsic motivation by fulfilling the three basic psychological needs of autonomy, relatedness, and competence, the relevant conclusions are based on students’ perceptions of formative assessment, and its specific implementation methods and effectiveness remain unclear. Therefore, in addition to focusing on motor skill indicators, this study also examines the three psychological indicators of autonomy, relatedness, and competence, aiming to explore the impact of different assessment methods on the fulfillment of basic psychological needs among beginner college students.

To this end, this study designed and implemented formative and summative assessments in a teaching context to explore the differential effects of these two assessment methods on college students’ tennis skills and the fulfillment of their basic psychological needs. Specifically, from a BPNT perspective, the study examines whether the ongoing process support inherent in formative assessment is more effective than the outcome-oriented summative assessment in meeting students’ needs for autonomy, relatedness, and competence, thereby promoting their skill acquisition. This study targets college tennis beginners and uses a randomized controlled experiment to test the following hypothesis: H1: After controlling baseline skill levels, students receiving formative assessment will demonstrate significantly better post-test performance in tennis skills than those receiving summative assessment.

H2: Following the course intervention, students who received formative assessment will demonstrate significantly higher levels of fulfillment of the basic psychological needs of autonomy, relatedness, and competence compared to students who received summative assessment.

## Methods

### Participants

This study was approved by the Institutional Review Board of the university (Approval No.: LL2024000191-FA). Between September 1 and 30, 2025, 128 undergraduate students (64 males, 64 females; *M*_age_ = 19.22, *SD*= 0.91) with no prior tennis experience were recruited. Participants were cluster-randomized to either a formative assessment group (*n*= 64) or a summative assessment group (*n*= 64). All completed a 10-week tennis course comprising one 90-minute session per week. Prior to the course, the researcher explained the study’s purpose, procedures, voluntary nature, and participant rights, addressing any questions. Subsequently, informed consent was obtained verbally with a show of hands, with all participants indicating their agreement to participate.

### Experimental design and procedure

#### Formative assessment course design

This study adopted the key strategies proposed by Black & Wiliam [13] to design a formative assessment intervention based on the three core elements of formative assessment: feed-up (where the learner is headed), feedback (the learner’s current position), and feed-forward (how the learner will reach the goal) as well as the three roles of teacher, peer, and learner. The intervention is divided into three phases: pre-class, in-class, and post-class. Pre-class: clarify individual learning objectives and success criteria. The instructor explains the success criteria for forehand and backhand techniques, which include specific qualitative and quantitative standards; then uploads videos of standard movements to the CANVAS platform (a school-specific online teaching platform). Students watch and learn independently, recording their reflections and setting personal learning goals in their individual journals. In class: process feedback and periodic assessments are used to determine the learner’s current progress. Feedback during class takes the form of both instructor feedback and peer feedback, both conducted in small groups. Instructor feedback involves providing guidance and suggestions to fixed groups during each class; peer feedback follows the “feedback sandwich” framework, which includes acknowledging the strengths of a peer’s practice, pointing out areas for improvement, and offering corresponding suggestions. Periodic assessments refer to skill tests conducted after the completion of basic tennis instruction. Post-class: actions to advance toward learning objectives. The instructor analyzes video recordings from the formative assessments to provide personalized feedback, while learners submit reflection journals to the CANVAS platform and engage in independent practice.

#### Summative assessment course design

The curriculum for the summative assessment group is also divided into three phases: pre-class, in-class, and post-class. Before class, each student receives a shared set of course objectives and a plan; during class, the instructor delivers a focused explanation of the content and provides demonstrations; after student practice, the instructor offers unified feedback on common issues; after class, students engage in independent practice without any form of feedback.

#### Procedure

Participants were cluster-randomized to either a formative assessment group or a summative assessment group. Participants in both groups attended a tennis forehand and backhand technique instruction course once a week for 90 minutes per session over a 10-week period. Each session consisted of a 15-minute warm-up, a 60-minute practice session, and a 15-minute cool-down. All instruction on forehand and backhand tennis techniques, as well as formative and summative assessments, were conducted by a single principal investigator. Each session included 32 participants and utilized eight adjacent tennis courts for instruction.

Prior to the experiment, all participants completed a pre-test of tennis stroke accuracy. The results showed no significant difference in baseline performance between the two groups (*p* > 0.05). Before the first session began, the instructor explained the criteria for successful forehand and backhand strokes to all participants and distributed the “technical guidelines and self-assessment checklist for forehand and backhand strokes” (Appendix A) to everyone, providing a standardized explanation of the technical points listed therein. This sheet served as an external cue for students to use during all subsequent practice sessions.

Research Procedure: The instructional experiment began with foundational forehand and backhand skills. Weeks 1–4 consisted of foundational instruction, weeks 6–9 of advanced instruction, and week 5 featured a mid-term assessment of tennis skills. A post-test of tennis skills was administered immediately following the conclusion of the 9th-week session, and a follow-up test was conducted in the 10th week. The Basic Psychological Needs Satisfaction Test is administered twice: the first time before the start of the teaching experiment, and the second time immediately after the post-test of tennis skills. All participants undergo pre-test, interim tests, post-tests, and follow-up tests for tennis skills; each test consists of 12 strokes with no feedback of any kind.

### Measurement instruments

#### Tennis testing

Tests were conducted in an open environment. To control confounding factors, a ball machine was used for both practice and testing. A camera recorded the entire process for later scoring. All tests were conducted on the same tennis court. The test utilized the “International Tennis Number (ITN) – Groundstroke Accuracy” protocol [52]. ITN is a globally recognized non-professional player rating system established by the International Tennis Federation (ITF) and has been widely adopted in China [53, 54]. As shown in Fig 1, the coach controlled the ball machine from point F (the center of the baseline), while the participant stood at point P (the center of the baseline). The machine served balls to the participant at point P at a speed of 40 km/h, with a 20-degree angle of elevation, at a rate of one ball every 3 seconds, aiming for a designated 50 cm × 50 cm area 50 cm beyond the opposite service line. The subject was required to complete 12 strokes, consisting of 6 forehand and 6 backhand shots, in the order of forehand followed by backhand. Scoring was determined based on the positions of the ball’s first and second bounces, with a total of 6 scoring zones ranging from 1 to 3 points; if the first bounce landed outside the scoring zones, 0 points were awarded; if the first bounce falls on the boundary line between two scoring zones, the higher-scoring zone is counted; if the second bounce occurs between the baseline and the bonus line, 1 bonus point is awarded; if it lands beyond the bonus line, double points are awarded. During the test, participants are required to hit the tennis ball into the valid scoring zones of the court to achieve the highest possible cumulative score, with a maximum total of 84 points.

**Fig 1.**
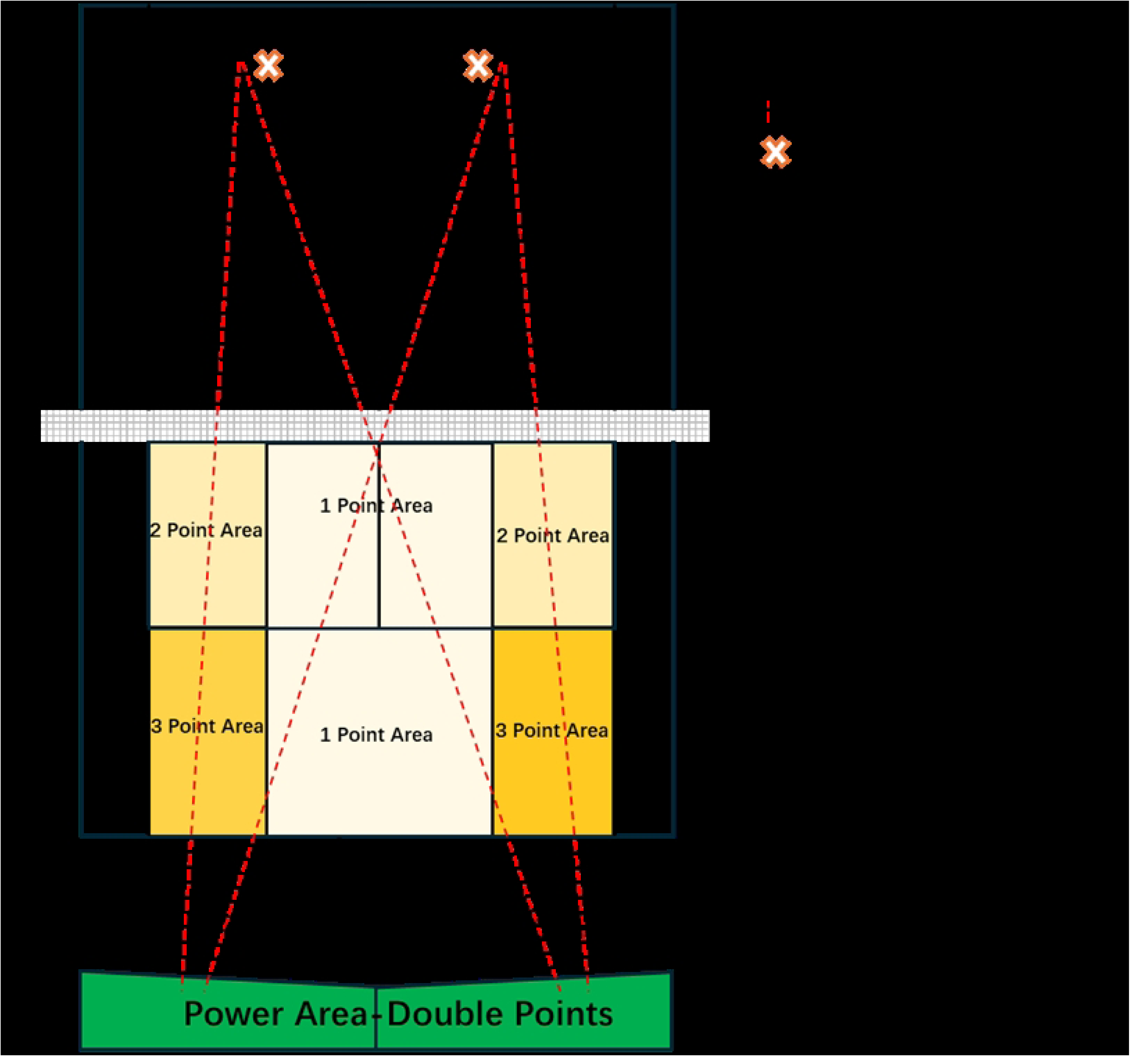
ITN—groundstroke accuracy.

### Basic Psychological Needs Satisfaction Scale (BPNSS)

The study utilized the Chinese version of the Basic Psychological Needs Satisfaction Scale (BPNSS), which was developed by Ryan & Deci [46] and translated, revised, and cross-culturally validated by Chen et al. [55]. This scale has been used in numerous studies to validate its cross-cultural and psychometric properties [56–58]. The scale uses the prompt “In physical education classes,….” It comprises 12 items that assess autonomy satisfaction (e.g., “In physical education classes, I have a sense of choice and freedom regarding what I do”), competence satisfaction (e.g., “In physical education classes, I feel capable of achieving my goals”), and relatedness satisfaction (e.g., “In physical education classes, I feel a sense of warmth when interacting with others”).The scale uses a 5-point Likert scale ranging from 1 (“Strongly Disagree”) to 5 (“Strongly Agree”), with higher scores indicating a higher degree of psychological need fulfillment. In this study, participants were required to complete the BPNS twice; the Cronbach’s alpha coefficients for the scale were 0.71 and 0.76, respectively.

### Statistical analysis

All data were analyzed using SPSS 26.0. Descriptive statistics were performed on participants’ gender, age, tennis skill scores at each time point, and scores on each dimension of psychological needs, reporting the mean (*M*) and standard deviation (*SD*).For the tennis accuracy test, a 2 × 4 mixed-effects ANOVA was conducted with evaluation type (formative assessment, summative assessment) and testing time (pre-test, mid-test, post-test, follow-up test), as well as their interaction, as independent variables, and tennis test scores as the dependent variable. Since the results of the Munchkin sphericity test were significant (*W* = 0.438, *p* < 0.001), violating the sphericity assumption, the F-values for all within-subject effects were reported using degrees of freedom corrected by the Greenhouse-Geisser method. For the Basic Psychological Needs Satisfaction Test, three 2×2 mixed-effects ANOVAs were conducted, with evaluation type (formative evaluation, summative evaluation) as the between-subjects variable and test timing (pre-test, post-test) and their interaction as the within-subjects variables, and autonomy, relatedness, and competence scores as the dependent variables, respectively. Pre- and post-test scores for all dimensions met the assumption of homogeneity of variances (Levine’s test, *p* > 0.05). Since the time factor included only two levels (pre-test and post-test), the sphericity assumption was automatically satisfied (*W* = 1.000); therefore, the results were presented and analyzed directly using the “assumed sphericity” approach. The significance level α was set at 0.05. In the ANOVA, the partial eta-squared (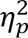) was used as the effect size measure to assess the practical importance of significant effects. According to Cohen’s (1988) criteria, 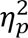 of ≥ 0.01 indicates a small effect, ≥ 0.06 a moderate effect, and ≥ 0.14 a large effect. When the results of the mixed-effects ANOVA indicated a significant interaction (Time × Group), a post-hoc analysis was conducted to clarify the specific differences between the two groups at different time points.

Prior to the intervention, there were no significant differences between the two groups in terms of tennis skills (*F* (1, 126) = 0.90, *p* = 0.345), autonomy (*t* (126) = 1.23, *p* = 0.220), relatedness (*t* (126) = 0.10, *p* = 0.922), and sense of competence (*t* (126) = 0.68, *p* = 0.500), meeting the homogeneity requirement.

## Results

### Descriptive statistics

The 128 undergraduate students were cluster-randomized to either a formative assessment group (*n* = 64) or a summative assessment group (*n* = 64), with a 50% male-to-female ratio in each group. The body mass index (BMI) was 22.57 ± 3.41 in the formative assessment group and 22.08 ± 4.03 in the summative evaluation group, with no significant difference between groups (*P* > 0.05).

### Tennis skill

Table 1 presents the tennis skill scores of participants in both groups at different time points and the results of the mixed-effects analysis of variance. The formative assessment group had higher average scores than the summative evaluation group at all time points, and scores in both groups showed an upward trend over time. The formative assessment group performed more notably during the post-test and follow-up test phases, and the gap between the two groups was greatest during the follow-up test phase. The results of the mixed-effects ANOVA indicated that the main effect of time was significant, *F* (1, 314, 165.595) = 1905.98, *p* < 0.001, 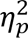 = 0.94. This suggests that the tennis skills of all participants improved significantly over time. The main effect of group was significant, *F* (1, 126) = 16.27, *p* < 0.001, 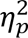 = 0.11, indicating that, when considering all four time points, the formative assessment group’s overall performance was significantly superior to that of the summative assessment group. The interaction between time and group was significant, *F* (1, 314, 165.595) = 32.93, *p* < 0.001, 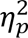= 0.21. This indicates that there was a significant difference in the patterns of skill performance over time between the two groups of participants.

**Table 1.** Mean scores for tennis skills and basic psychological needs, and results of mixed-effects analysis of variance.

| Scales and<br>measurements | FA group<br>(*n*= 64) | SA group<br>(*n*= 64) |  | Group | Time | Group*Time<br>interaction |
| --- | --- | --- | --- | --- | --- | --- |
|  | mean (*SD*) | mean (*SD*) |  |  |  |  |
| **Tennis** |  |  |  |  |  |  |
| Pre-test | 6.59 ± 2.16 | 6.27 ± 1.74 | ***F*** | 16.27 | 1905.98 | 32.93 |
| Mid-test | 15.64 ± 3.21 | 14.56 ± 3.57 | ***p*** | < 0.001 | < 0.001 | < 0.001 |
| Post-test | 30.56 ± 6.05 | 27.06 ± 6.51 | **$\eta_p^2$** | 0.11 | 0.94 | 0.21 |
| Follow-up (1<sup>st</sup><br>week) | 32.61 ± 6.15 | 25.80 ± 6.12 |  |  |  |  |
| **BPNS-Autonomy** |  |  |  |  |  |  |
| Pre-test | 14.25 ± 2.17 | 13.80 ± 1.99 | ***F*** | 18.97 | 168.28 | 148.93 |
| Post-test | 16.30 ± 1.50 | 13.86 ± 2.02 | ***p*** | < 0.001 | < 0.001 | < 0.001 |
|  |  |  | **$\eta_p^2$** | 0.13 | 0.57 | 0.54 |
| **BPNS-Relatedness** |  |  |  |  |  |  |
| Pre-test | 13.94 ± 1.59 | 13.91 ± 1.97 | ***F*** | 4.69 | 94.26 | 78.41 |
| Post-test | 15.30 ± 1.64 | 13.97 ± 2.05 | *p* | 0.032 | < 0.001 | < 0.001 |
| | | | $\eta_p^2$ | 0.04 | 0.43 | 0.38 |
| **BPNS-Competence** |  |  |  |  |  |  |
| Pre-test | 13.50 ± 2.10 | 15.77 ± 2.02 | *F* | 8.41 | 179.87 | 44.05 |
| Post-test | 13.25 ± 2.09 | 14.02 ± 2.00 | *p* | 0.004 | < 0.001 | < 0.001 |
| | | | $\eta_p^2$ | 0.06 | 0.59 | 0.26 |

*F* =Two-way mixed-effects ANOVA; FA: formative assessment; SA: summative assessment.

Further simple effects analysis indicated that, in within-group comparisons, the formative assessment group showed significant improvement across all consecutive time points (*p* < 0.001). Notably, during the follow-up phase following the intervention, their performance remained significantly higher than that at the post-test, with an *MD* of 2.05, 95% *CI* [1.649, 2.444], *p* < 0.001.For the summative evaluation group, although progress from the pre-test to the post-test was significant (*p* < 0.001), performance declined significantly from the post-test to the follow-up phase, with an *MD* of −1.27, 95% *CI* [−1.663, −0.868], *p* < 0.001.In the between-group comparison (see Fig 2), there was no significant difference between the two groups at the pre-test, *MD* = 0.33, 95% *CI* [−0.357, 1.014], *p* = 0.345, indicating homogeneity in pre-test levels. At the mid-term assessment, the difference between the two groups was also not significant, *MD* = 1.08, 95% *CI* [−0.110, 2.266], *p* =0.075.At the post-test, the formative assessment group significantly outperformed the summative assessment group, *MD* = 3.50, 95% *CI* [1.303, 5.697], *p* = 0.002.In the follow-up test, the difference between the groups reached its maximum and was highly significant, with an *MD* of 6.81, a 95% *CI* of [4.667, 8.958], and a *p*-value of < 0.001. Fig 2 illustrates the different trajectories of performance between the two groups. The two curves were close at the pre-test and rose in tandem thereafter but diverged markedly from the post-test through the follow-up phase: the curve for the formative assessment group continued to rise, while the curve for the summative assessment group flattened and declined slightly. This result indicates that the formative assessment group’s intervention was more effective in promoting the long-term retention of tennis skills among college students.

**Fig 2.**
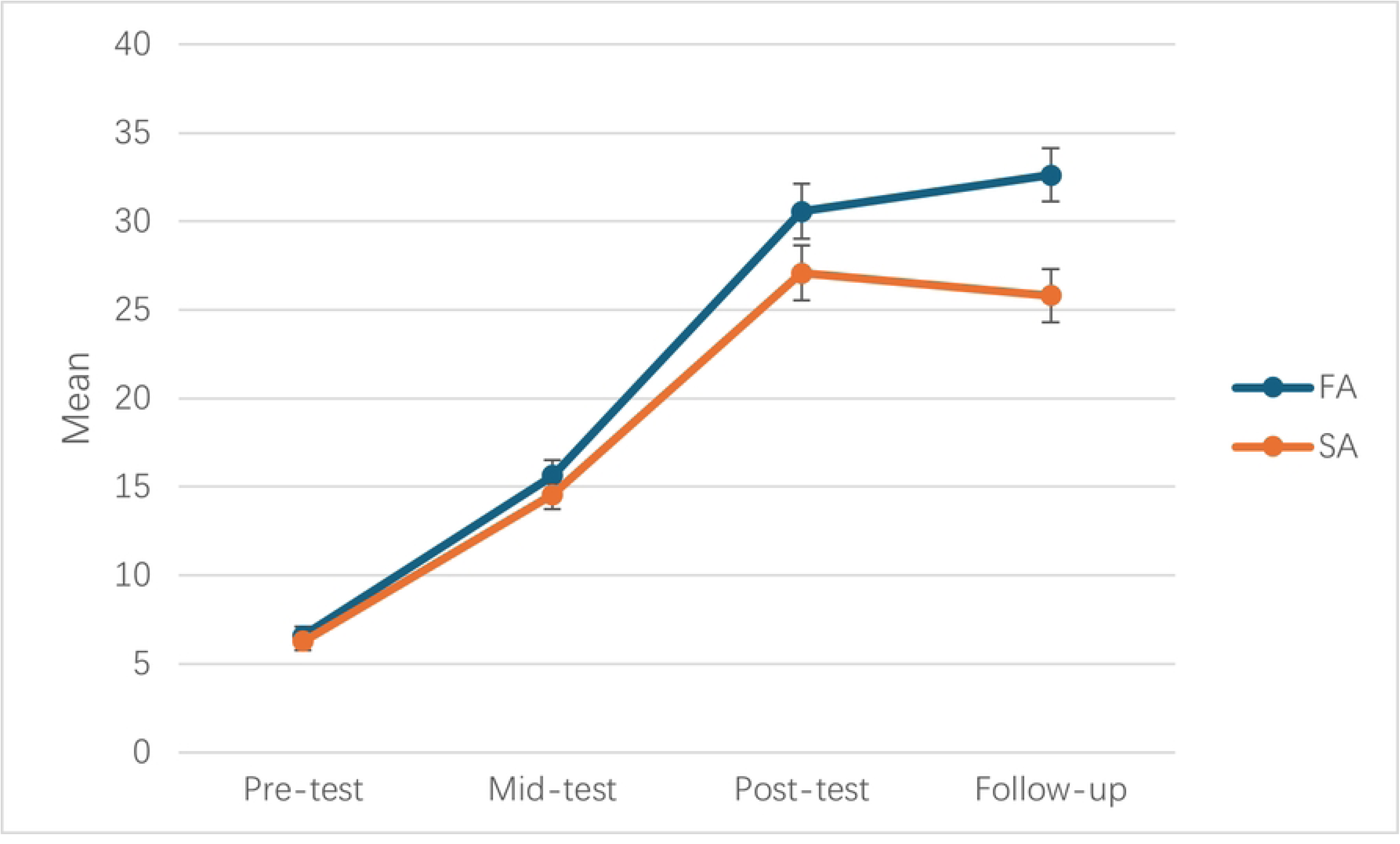
Simple slope analysis of the group-time interaction effect on tennis skills.

### Basic psychological needs satisfaction

Table 1 presents the scores for the satisfaction of basic psychological needs at different time points and the results of the mixed-effects analysis of variance for the two groups of participants. The results indicate that, compared to summative evaluation, formative evaluation is more effective in enhancing college students’ autonomy, relatedness, and competence.

The results of the ANOVA for autonomy showed a significant main effect of time, *F*(1, 126) = 168.28, *p* < 0.001, 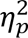 = 0.57; a significant main effect of group, *F*(1, 126) = 18.97, *p* < 0.001, 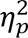 = 0.13;The interaction between time and group was significant, *F*(1, 126) = 148.93, *p* < 0.001, 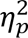 = 0.54. Further simple effects analysis revealed that, in within-group comparisons, the post-test scores in the formative assessment group were significantly higher than the pre-test scores, *MD* = −2.05, 95% *CI* [−2.274, −1.819], *p* < 0.001; whereas there was no significant difference in pre- and post-test scores for the summative evaluation group, *MD* = −0.062, 95% *CI* [−0.290, 0.165], *p* = 0.588.In the between-group comparison (see Fig 3), there was no significant difference in scores between the two groups at the pre-test, *MD* = 0.45, 95% *CI* [−0.274, 1.180], *p* = 0.220;at the post-test, the scores of the formative assessment group were significantly higher than those of the summative assessment group, *MD* = 2.44, 95% *CI* [1.816, 3.059], *p* < 0.001. The results indicate that formative assessment interventions can significantly enhance students’ autonomy, whereas traditional summative assessment does not produce this effect.

**Fig 3.**
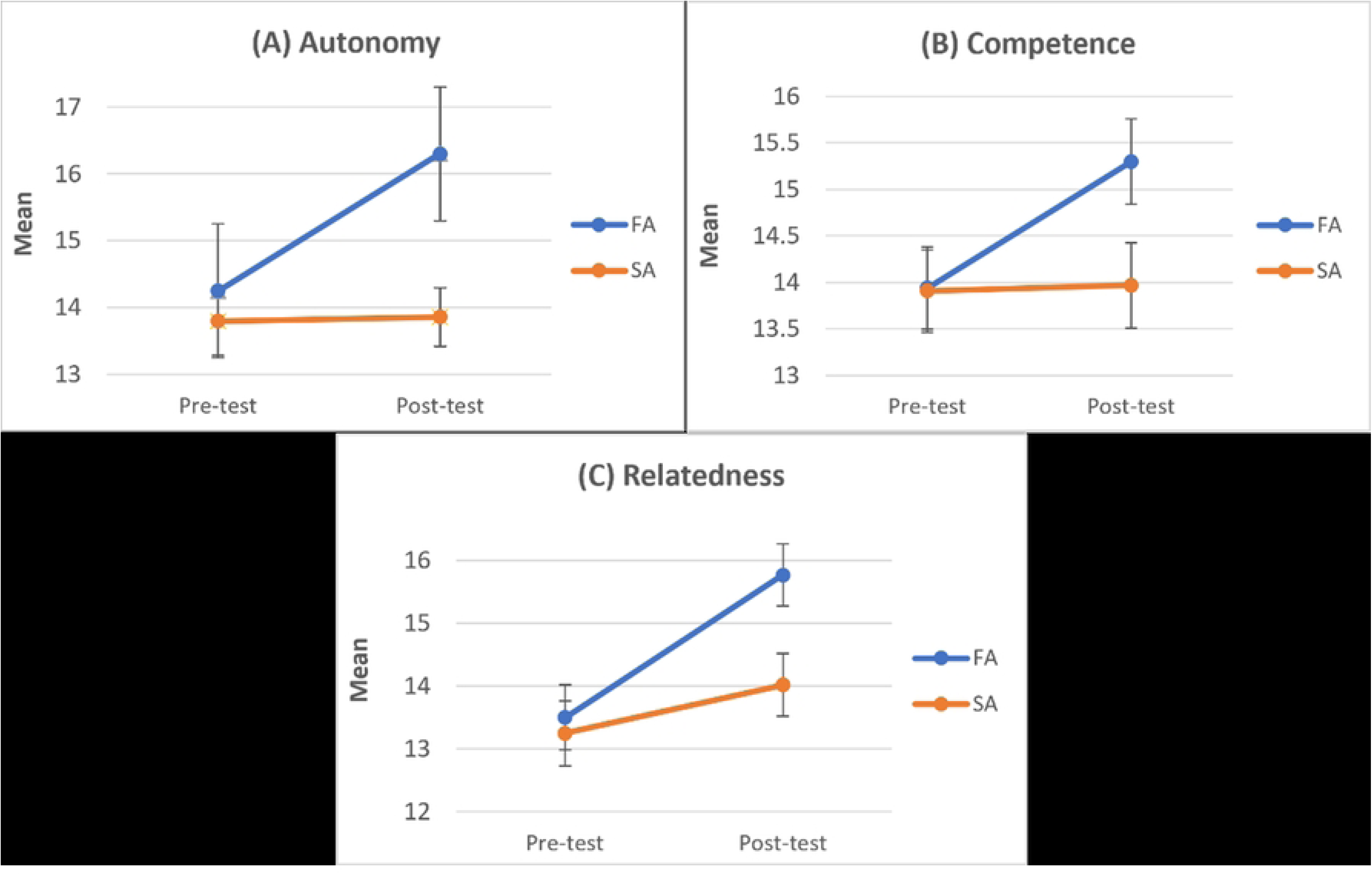
Simple slope analysis of the group-time interaction effect on basic psychological needs.

The results of the ANOVA for relatedness showed a significant main effect of time, *F*(1, 126) = 94.26, *p* < 0.001, 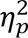 = 0.43; the main effect of group was significant, *F*(1, 126) = 4.69, *p* = 0.032, 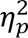 = 0.04;The interaction between time and group was also significant, *F*(1, 126) = 78.41, *p* < 0.001, 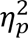 = 0.38, with a high effect size. Further simple effects analysis revealed that, within-group comparisons showed the post-test scores in the formative assessment group were significantly higher than the pre-test scores, *MD* = −1.36, 95% *CI* [−1.564, −1.154], *p* < 0.001; whereas there was no significant difference between pre- and post-test scores in the summative assessment group, *MD* = −0.06, 95% *CI* [−0.267, 0.142], *p* = 0.547. In between-group comparisons (see Fig 3), at the pre-test, there was no significant difference in relational scores between the two groups, *MD* = 0.03, 95% *CI* [−0.596, 0.659], *p* =0.922; however, at the post-test, the formative assessment group’s scores were significantly higher than those of the summative assessment group, *MD* = 1.33, 95% *CI* [0.679, 1.977], *p* < 0.001. The results indicate that formative assessment interventions significantly promote students’ relational skills, whereas summative assessment methods have no significant effect on this dimension.

The results of the ANOVA for competence showed a significant main effect of time, *F*(1, 126) = 179.87, *p* < 0.001, 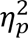 = 0.59; a significant main effect of group, *F*(1, 126) = 8.41, *p* = 0.004, 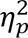 = 0.06;The interaction between time and group was also significant, *F*(1, 126) = 44.05, *p* < 0.001, 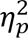 = 0.26, with a high effect size. Further simple effects analysis revealed that, in within-group comparisons, the post-test scores in the formative assessment group were significantly higher than the pre-test scores, *MD* = −2.27, 95% *CI* [−2.582, −1.949], *p* < 0.001;and the post-test scores in the summative evaluation group were also significantly higher than the pre-test scores, *MD* = −0.77, 95% *CI* [−1.082, −0.449], *p* < 0.001.In the between-group comparison (see Fig 3), there was no significant difference in scores between the two groups at the pre-test, *MD* = 0.25, 95% *CI* [−0.482, 0.982], *p* = 0.500; however, at the post-test, the scores of the formative assessment group were significantly higher than those of the summative assessment group, *MD* = 1.75, 95% *CI* [1.046, 2.454], *p* < 0.001. The results indicate that both assessment methods can enhance college students’ competence, but the formative assessment intervention is more effective.

## Discussion

This study used a randomized controlled trial to evaluate the effectiveness of formative and summative assessment strategies in improving college students’ tennis performance. The findings demonstrate that, in addition to improving skill acquisition, formative assessment was more effective than summative assessment in satisfying the students’ basic psychological needs of autonomy, relatedness, and competence. By providing clear learning objectives, continuous process feedback, and personalized guidance, formative assessment supported sustained skill development and greater psychological engagement. These results suggest that incorporating personalized goals, process-oriented evaluation, and tailored feedback into tennis instruction could promote both skill development and positive psychological outcomes in college physical education.

The results suggest that both formative and summative assessments significantly improved college students’ tennis performance. This is consistent with the findings of [17, 27], which suggest that clear-goal, structured assessment strategies can enhance learners’ athletic performance. Both types of assessment serve as effective tools for providing evidence of learning to inform instructional decisions, and instructors should foster synergy rather than conflict between them [59].

Specifically, in teaching practice, both assessment courses employed a structured, goal-oriented instructional designs in teaching practice to ensure that all participants received an equal number of class hours. This approach maintained consistency in the overall course objectives, basic structure, and volume of practice. Secondly, feedback was provided by professional instructors with extensive teaching experience in the field. And this feedback was aligned with the instructional objectives. As Black & Wiliam [13]and Hattie [60] have noted, feedback with high density of information has a significant impact on learners’ cognitive and motor skill outcomes. This feedback could take various forms, such as video, audio, and was primarily intended to provide learners with cues or reinforcement. In teaching practice, the teacher’ feedback in both assessment courses consistently adhered to the overall instructional objectives. Furthermore, since the issues faced by novice learners are relatively universal, such as, the completeness of stroke mechanics, the position of the contact point, and the racket face angle during the stroke, the teacher’ feedback served as both a prompt and a reinforcement to enhance the tennis skills of the college students. Thirdly, all participants were college students with high cognitive ability and good physical fitness. These characteristics facilitated their reception and execution of task feedback, which may have optimized the intervention’ overall effectiveness. Therefore, this study demonstrates that structured summative assessment can still serve as a foundational tool for organizing instruction and evaluating short-term learning outcomes, even under constraints of limited teaching resources or time.

The results demonstrate that the formative assessment group showed significantly better long-term skill retention and continued development than the summative assessment group. An analysis of interaction clearly revealed distinct learning trajectories between the two groups: the formative assessment group continued to improve from the post-test to the follow-up test, while the summative assessment group showed a flattening trend or signs of decline. This suggests that formative assessment is more effective than summative assessment in the follow-up phase of skills development. This finding is consistent with the results of studies by Barrientos Hernán et al. [27] and Ní Chróinín & Cosgrave [17], which suggest that formative assessment yields can significantly improve learning outcomes by closing the gap between learners’ “current performance” and “success criteria” [12, 61]. As this study demonstrates, formative assessment is more effective than summative assessment in promoting the follow-up and internalization of learners’ tennis skills. This may be due to the fact that formative assessment systematically integrates the three core functions of feedback: feedforward (where the learner is headed), feedback (where the learner is currently positioned), and feedback (how the learner will reach the goal) [15, 61, 62]. This finding strongly supports the theoretical hypothesis that formative assessment optimizes athletic performance and promotes deep learning.

Specifically, the formative assessment intervention strategy in this study is implemented across three phases, before, during, and after class, through the multi-role participation of teachers, peers, and the learners themselves.

In the pre-class phase, the intervention focuses on clarifying learning objectives and establishing success criteria. The students need to design personalized, specific goals based on the course’s overall objectives. Then students and teachers jointly discuss the differences between individual learning goals and overall course objectives, which further helps the students understand the course goals and success criteria.

During the in-class phase, through continuous process feedback provided by teachers and peers, learners can monitor and adjust their learning process in real time. In practice, teachers provide targeted guidance and suggestions to fixed groups during each class session. Peer feedback employs the “sandwich method,” which includes an assessment of performance and relevant suggestions for improvement. This exercise enhances student engagement in the classroom and encourages students to take ownership of their learning. However, this study also observed that the implementation and quality of peer feedback are highly dependent on the feedback literacy of both the provider and the recipient. To enhance the quality of peer feedback during the intervention and compensate for the participants’ lack of tennis expertise, this study introduced a peer feedback scaffold: the “technical guidelines and self-assessment checklist for forehand and backhand strokes” (Appendix A), which incorporates clear criteria for technical success.

In the post-class phase, this study emphasized reflection and self-directed practice based on personalized feedback, encouraging learners to translate external feedback into concrete improvement actions. In practice, instructors used smartphones or iPads to record students’ technical movements and provided personalized feedback through features such as slow-motion playback, angle adjustments, and still images, helping students identify key errors and optimize their execution. Student feedback indicates that this feedback has motivated them to practice outside of class. This finding is consistent with the research by [28–30], which suggests that the use of tools such as digital video in the classroom can significantly enhance student motivation and classroom engagement, and directly influence athletic performance.

In summary, through the three phases of pre-class, in-class, and post-class, learners receive continuous, goal-oriented process feedback. These specific instructions and partial solutions may effectively reduce the cognitive load on beginners during the acquisition of complex skills, thereby serving as cognitive scaffolding. Consequently, as learners’ abilities improve, this external scaffolding is gradually internalized, supporting the follow-up and transfer of their tennis skills.

However, summative assessment, due to its inherent “uniform and standardized” nature, focuses on the final judgment of learning outcomes rather than providing continuous support for the learning process. While this assessment method may drive performance in the short term, its lack of continuous responsiveness to the individual learner’s progress may be the primary reason why motor skills decline once the assessment concludes [15].

The results of this study indicate that formative assessment can significantly enhance college students’ levels of fulfillment across three dimensions: autonomy, relatedness, and competence. This is consistent with the findings of Herrero-González et al. [33] and Leenknecht et al. [14], which suggest that formative assessment influences students’ intrinsic motivation by affecting their levels of fulfillment regarding autonomy, relatedness, and competence. The implementation of formative assessment is closely linked to the theory of basic psychological needs. For example, setting personalized learning goals and maintaining reflection journals support autonomy; providing continuous process feedback enhances competence; and fostering peer interaction satisfies relational needs. Together, these methods promote the fulfillment of basic psychological needs. According to Self-Determination Theory [47, 63], an individual’s level of motivation depends on whether their three basic psychological needs—autonomy, relatedness, and competence are sufficiently met. When students perceive interest or meaning in their task engagement, their extrinsic motivation gradually internalizes, leading to an intrinsic goal orientation [45, 48], thereby increasing the possibilities of the positive learning outcome and long-term skill follow-up.

Specifically, the results of this study indicate that formative assessment exerts a unique “catalytic effect” on autonomy and relatedness. Significant improvements were observed only in the formative assessment group. And a “reinforcing effect” on competence, which means that the magnitude of improvement in the formative assessment group was significantly greater than that in the summative assessment group.

Firstly, regarding autonomy, formative assessment empowers students by introducing self-assessment and peer assessment, and by allowing them to independently set improvement goals and practice pathways based on teacher feedback.

This stands in stark contrast to the one-way, uniform evaluation model of summative assessment. When students experience autonomy in decision-making and choice during the acquisition of tennis skills (such as adjusting the kinetic chain for a forehand stroke), the “self-determination” component of their intrinsic motivation is strengthened.

Secondly, regarding relational aspects, the collaborative learning activities embedded in formative assessment courses, which include intra-group observation and feedback, as well as personalized post-class feedback from teachers, foster a supportive classroom atmosphere centered on learning and development. In this process, students not only engage in dialogue with teachers regarding skill mastery but also establish mutually supportive learning connections with peers. These steps fulfill students’ need for positive social relationships. Traditional summative assessment, however, being a uniform and standardized evaluation, struggles to provide these critical psychological nutrients.

Finally, regarding a sense of competence, although both assessment methods enhance students’ sense of competence, the “reinforcement effect” of formative assessment is particularly pronounced. Specifically, tennis is an open-ended motor skill, and its effective learning and mastery present unique pedagogical challenges. Open-ended skills require learners to execute technical movements quickly and in a coordinated manner based on real-time information within dynamic, unpredictable environments (such as a peer’s return, wind direction, and court conditions).Successful mastery of tennis techniques, such as forehand and backhand strokes, relies heavily on the coordination of the kinetic chain (from lower-body push-off and trunk rotation to the transfer of power through the racket swing), as well as the continuous perception, correction, and optimization of fine motor parameters such as timing and racket face control [64, 65]. Formative assessment courses use methods such as video analysis, periodic testing, and personalized feedback to direct students’ attention toward specific areas for improvement, rather than focusing solely on the outcome of the shot. This feedback, which uses “mastery” and “progress” as benchmarks, helps students develop a realistic and positive sense of competence. Especially for students with lower initial skill levels, it effectively mitigates the frustration caused by poor early performance, thereby more effectively fostering the belief that “I am capable of achieving my goals.” In contrast, summative assessment courses fall short in providing more specific and personalized information regarding student efficacy.

### Practical Value

This study indicates that both formative and summative assessments can significantly improve college students’ tennis performance. Compared to summative assessment, formative assessment features clear learning objectives and provides continuous feedback during the learning process. It is more effective at promoting the long-term retention of tennis skills and meeting students’ psychological needs for autonomy, relatedness and competence. Therefore, this study’s practical significance lies in the following: in alignment with a clearly defined instructional framework, teachers should appropriately integrate these two assessment methods, enabling them to function synergistically and inform pedagogical decision-making. When evaluating the effectiveness of physical education teaching strategies, the focus should not be limited to immediate or short-term outcomes, but should also consider long-term retention of motor skills and deep learning. This helps students consolidate the skills they have learned and enhances their ability to apply them in practice. Furthermore, this study found that meeting students’ psychological needs for autonomy, relatedness, and competence is central to effective teaching strategies. Creating a positive socio-psychological environment can stimulate students’ intrinsic motivation to learn and their willingness to participate consistently.

### Limitations and Future Directions

This study provides important practical evidence for the field of physical education, particularly with regard to the impact of formative and summative assessments on college students’ tennis performance and basic psychological needs of them. However, certain limitations remain. Firstly, in terms of research scope, this study focused on tennis; future research could extend to open-skill sports such as badminton and table tennis, and include participants of different abilities to examine whether formative assessment can be generalized across different types of sports and stages of skill development. Secondly, the digital tools used for assessment and feedback in this study still present issues such as delayed processing of feedback and reliance on teachers’ subjective experience. Future efforts should explore the ways to apply digital formative assessment tools that enable more timely feedback and enhance the objectivity and teaching effectiveness of assessments.

### Conclusion

This study confirmed the effectiveness of both formative and summative assessment in improving college students’ tennis performance. Specifically, formative assessment was proved to be more effective in promoting the long-term retention of skills and in satisfying students’ basic psychological needs, such as autonomy, relatedness, and competence, through the provision of clear learning objectives and continuous process feedback. The results suggested that physical education instruction should incorporate formative assessment elements, such as personalized goals, process-oriented evaluation, and individualized feedback, in order to support long-term motor skill development, enhance student motivation and engagement, and addressing these psychological needs. Future research could expand to include a wider range of open-skill sports and participants of varying skill levels. It could also explore the use of digital formative assessment tools to improve the timeliness and objectivity of feedback, thereby optimizing instructional effectiveness further.

## Supporting information

Supplementary file: technical guidelines and self-assessment checklist for forehand and backhand strokes (Appendix A)

## Data Availability

All relevant data are within the manuscript and its supporting information files.

## Acknowledgments

We would like to thank all the authors for their contributions for this manuscript, and all participants for participating in this study.

## Author Contributions

Conceptualization: Yazhuo Wang.

Data curation: Yazhuo Wang.

Project administration: Yazhuo Wang.

Writing – original draft: Yazhuo Wang, Yan Luo.

Writing – review & editing: Yazhuo Wang, Yan Luo.

All authors read and approved the final manuscript.

